# Dual Guidance in Regional Anesthesia – Influence of Needle Electrode Configuration on Stimulation Success at Sciatic Nerve; A Randomized, Controlled Pilot Trial

**DOI:** 10.1101/2022.03.01.22271677

**Authors:** Julia Wegner, Martin Ertmer, Sascha Tafelski, Edda Klotz, Jürgen Birnbaum

## Abstract

**Introduction:** In contrast to ultrasound technology (US), peripheral nerve stimulation (PNS) for regional anesthesia was little improved in recent years. When using the combination of both techniques, PNS can give additional information for nerve localization to improve safety and success of regional anesthesia. There are influencing factors on the success rate of stimulation in PNS remaining uninvestigated in a clinical setting to date. This randomized controlled pilot trial evaluates the impact of shape and size of stimulation needles electrodes under dual guidance conditions.

**Methods:** In a randomized controlled clinical trial 35 participants undergoing lower limb surgery received a preoperative proximal sciatic nerve block in dual guidance technique. Use of facet needles with point shaped electrodes (N=19, facet group) were compared with tuohy needles with large electroconductive tips (N=16, touhy group). Stimulation success at minimal distance between needle tip and nerve was recorded. Block success and complications of regional anesthesia were assessed.

**Results:** In 87% of successful stimulation (20 of 23) an ultrasound-proven contact of needle tip and sciatic nerve was necessary to elicit a motor response. More successful stimulations could performed using facet needles (84%, 16/19) compared to tuohy needles (44%, 7/16, p=0.03). If stimulation was successful the number of successful sensory blockades was increased (78%, 18/23, p=0.02). No serious complications of regional anesthesia were recorded.

**Discussion:** This pilot trial suggests that stimulation needles with small electrodes may be more reliable in indicating a contact of needle and nerve, which may improve safety and success of proximal sciatic nerve blocks.

## INTRODUCTION

In recent years, ultrasound (US) has made decisive advances in regional anesthesia and safety has been increased further.^1^ Compared to nerve stimulation, fewer vascular punctures occur and the risk of local anesthetic intoxication (LAST) is reduced.^2; 3^ Certain blockades are experiencing a renaissance with ultrasound, like for example the supraclavicular block and others are becoming safer, like the interscalene block with a reduced incidence of phrenic nerve block.^4^

Peripheral nerve stimulation (PNS) has been established as a regularly used clinical procedure several decades ago.^5^ Experimental studies showed an equal precision in needle placement of PNS compared to ultrasound while clinical trials investigated a similar success rate and comparable incidence of complication in axillary plexus block.^6; 7^ PNS is a useful tool to identify nerval structures in poor sonographic conditions.^8^ The combination of both techniques was the advocated approach for many physicians.^9^ Vassiliou et al. showed a reduction in intraneural injection and hematoma when using ultrasound and nerve stimulation simultaneously.^6^

While US has experienced several technical improvements, there are influencing factors on the stimulation success in PNS remaining unclear. Recommendations for current intensity differ between studies and there are different methods on how to deal with a stimulation response.^10; 11^

Additionally the specific properties of the target nerve have an influence on the stimulation success. The sciatic nerve, as a frequently investigated example, has a strong connective tissue sheath. This paraneural layer, which acts as an anatomical barrier as well as chemical and physical insulation can block stimulation current and local anesthetics.^12; 13^ Recent investigations also suggest that ultrasound imaging of the sciatic nerve at infragluteal access is worse compared to other peripheral nerves.^14^ In these cases PNS could give important additional information for nerve localization to increase patient safety.

The importance of technical equipment for PNS was little considered so far. Simulations suggested that different configurations and sizes of the electrodes at the needle tip lead to differences in the resulting electrical fields.^15; 16^ This clinical study investigates, whether the differently sized and shaped uninsulated electrodes of the needle tips have an impact on the stimulation success.

## METHODS

The study was approved by the ethics committee of Charité Universitätsmedizin Berlin with application number EA1/248/17. All data were collected at Charité Universitätsmedizin Berlin, Campus Charité Mitte, and the trial is registered with NCT03383770 at ClinicalTrials.gov (https://clinicaltrials.gov/ct2/show/NCT03383770?cond=NCT03383770&draw=2&rank=1). Participants were enrolled from February 2018 until September 2019.

This randomized controlled trial was designed to evaluate the effect of differences in size and shape of stimulating needles electrodes on stimulating success of the sciatic nerve. Therefore, tuohy needles (Arrow StimuCath 2, 18G, Teleflex, Wayne, USA) with large stimulation surfaces and facet needles (Uniplex 19G, Pajunk GmbH, Geisingen, Germany) with small, point shaped electrodes were compared (Figure 1). Both regional anesthesia cannulae were approved for the use in regional blockades in humans.

**Figure 1.**
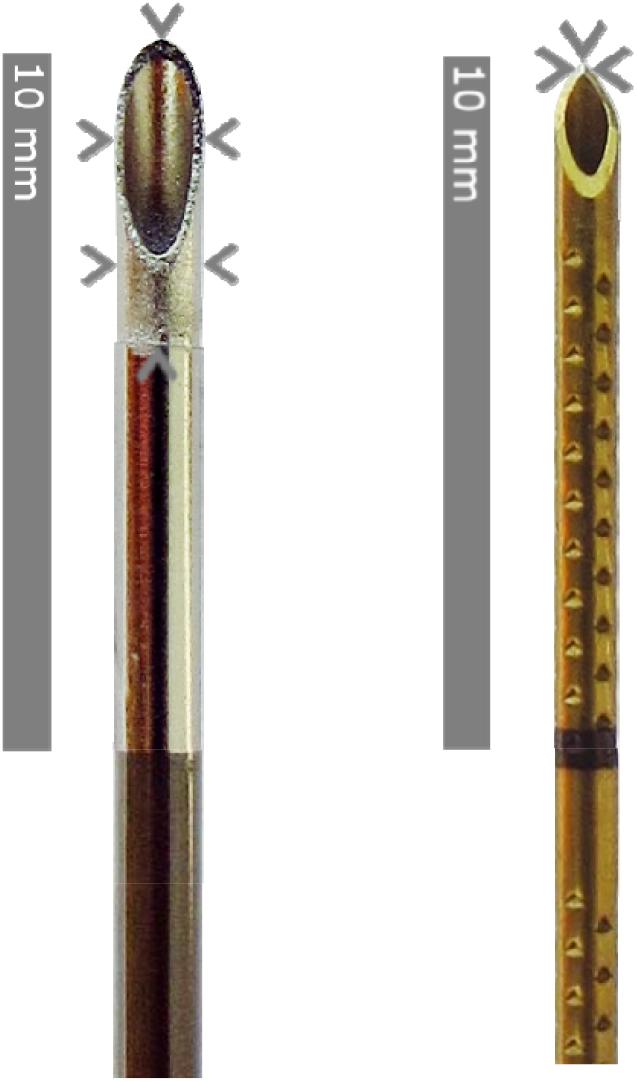
Comparison of Tuohy needle (left, Arrow StimuCath 2, 18G, Teleflex, Wayne, USA) and Facet needle (right, Uniplex 19G, Pajunk GmbH, Geisingen, Deutschland) with arrow heads marking uninsulated electrodes

Patients undergoing orthopedic surgery of the lower limb provided written consent and were assessed for eligibility. Participants had to be ≥ 18 years of age and did not attend another interventional study simultaneously. Exclusion criteria were pregnancy, preexisting nerve damages in the target area, diabetes mellitus, alcohol dependency, allergy to local anesthetics, implanted pacemaker, additionally planed spinal anesthesia and poor ultrasound visibility at the beginning of the block performance (VIS>2) as previously published by Birnbaum et al.^14^

Participants were randomized equally into two groups using randomization software following a block randomization list allowing an interim analysis after 10 participants each. In facet group the Uniplex needles were used and in the tuohy group the blocks were performed with needles from Arrow StimuCath 2 set. While the treating physicians were unblinded, participants were blinded to group assignment.

### Interventions

In both groups sciatic nerve blockades via infragluteal access were performed in dual guidance technique by the same experienced physician (J.B.) in out of plane technique.

Participants were awake and not sedated to be able to report about pain and dysesthesia. The sciatic nerve localizations were sonographically assessed using an ultrasound device M-Turbo^®^ and convex probe (C60x/5-2 MHz Transducer, FUJIFILM SonoSite, Inc. Bothell, USA). After femoral and sciatic tuberositas had been localized the tendon insertion of ischiocrural muscles was followed distally. Sciatic nerve was found laterally of this point. In slim participants a linear probe (HFL50x/15-6 MHz Transducer, FUJIFILM SonoSite, Inc. Bothell, USA) was used. Before each block was performed, the target structures were identified and classified with a Visibility score (VIS). Only participants providing VIS 1 (“all structures are detected reliable immediately or after a few corrections of the ultrasound probe position”^14^) and 2 (“the target structure detected reliable only after several corrections of the ultrasound probe”^14^) were included in this trial. The nerve stimulator (MultiStim SWITCH, Pajunk GmbH, Geisingen, Germany) was set with 2.0mA, 2Hz and 0.1ms as initial settings.

Puncture sites were set in sterile conditions and Lidocain 1% was injected superficially without affecting deeper tissue to not influence stimulation response. Under dual guidance the needle tip was moved towards the sciatic nerve until a motor response of the tibial or common peroneal nerve region could be noticed. The resulting distance between needle and nerve was recorded. Then the current was reduced by 0.5 mA steps gradually and the needle was moved further. If a motor response reappeared, the distance was recorded again and the procedure repeated until minimal current settings of 0.5 mA. Without motor response the needle tip was moved forward until sonography revealed direct contact between needle tip and nerve.

The final position of the needle tip was either found at minimal distance to the sciatic nerve with preserved motor response or at direct contact between needle tip and nerve without any stimulation success. After establishing the final position of the needle tip the tissue around the sciatic nerve was dissected with injection of glucose 5%. Finally, 20 ml Ropivacaine 0.75% were injected dorsal of the nerve and the needle was removed. Before surgery all patients without contraindications received an additional block of the saphenous nerve as well as general anesthesia.

### Outcome assessment

The inital outcome was the ultrasound-based measurement of the distance between needle tip and sciatic nerve which was necessary for successful stimulation at minimal current.

During the study it became apparent that a measurement of the distance between needle tip and nerve was infeasible because there was nearly no stimulation success without direct contact. Therefore the primary outcome was adjusted to be the proportion of successful nerve stimulations upon direct contact between needle tip and nerve.

As secondary outcome the effectiveness of the sensory and motor blockades depending on the preceding success of PNS were recorded. The effectiveness was assessed in 15-minute intervals after the performance of the block until general anesthesia was initiated and once in the recovery room after surgery. Success of motor blockade was measured by dorsal extension and plantar flexion of the foot and toes and sensory blockade was assessed by temperature and contact sensibility tested on the back of the foot. Paresthesia and complications of regional anesthesia were recorded.

### Statistical analyses

A sample size calculation could not be performed beforehand due to a lack of preexisting data comparing the stimulation success of tuohy and facet needles. Therefore we chose to perform a pilot trial consisting of 40 participants. Those meeting no exclusion criteria at the time of needle placement were included in statistical analyses performed with IBM SPSS Statistics 24. Depending on scale level and distribution, data were analyzed using a chi-square, Student’s t-test and Mann-Whitney-U-test, as appropriate. Normal distribution was analyzed using a Kolmogorov-Smirnov-test and graphical analyses. A sensitivity analysis using binary logistic regression as well as bootstrap analysis with a Cochran-Mantel-Haenszel test was performed to exclude influence of demographic differences between groups on primary endpoint. With the help of a Hosmer-Lemeshow-test the model quality of regression analysis could be verified. For all analyses, a two-sides p-value of <0.05 was defined as statistical significance level.

## RESULTS

### Study population

Altogether, 102 Participants were assessed with 40 consenting participants matching the eligibility criteria who were randomized to facet group (N=20) or tuohy group (N=20) either (figure 2). Two participants, one in each group, had to be excluded after surgery was rescheduled. Additionally, in tuohy group three participants were excluded from the analyses due to new reported exclusion criteria prior to needle placement. The full analysis set included 35 participants (facet group, n=19; tuohy group, n=16). Demographic characteristics for this subgroup are itemized in table 1. Participants differed significantly in age between the two groups. In average participants in facet group were about 8 years younger than in tuohy group with a total range from 47 to 84 years of age. A majority of participants in both groups were obese at a mean BMI of 29.1kg/m^2^. The most frequently performed intervention was total knee arthroplasty.

**Table 1.**
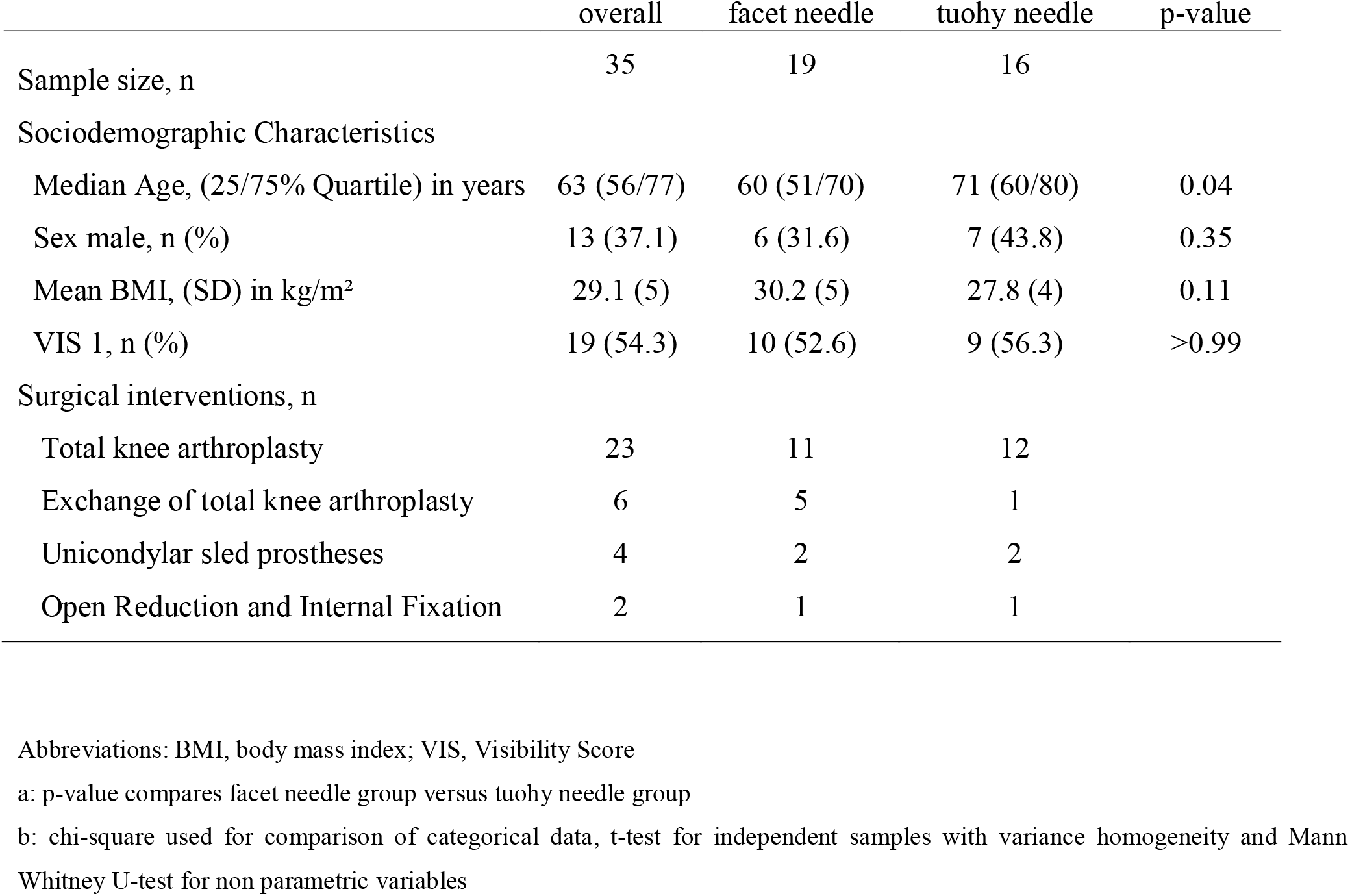
Demographics in full analysis set.

|  | overall | facet needle | tuohy needle | p-value |
| --- | --- | --- | --- | --- |
| Sample size, n | 35 | 19 | 16 |  |
| Sociodemographic Characteristics |  |  |  |  |
| Median Age, (25/75% Quartile) in years | 63 (56/77) | 60 (51/70) | 71 (60/80) | 0.04 |
| Sex male, n (%) | 13 (37.1) | 6 (31.6) | 7 (43.8) | 0.35 |
| Mean BMI, (SD) in kg/m<sup>2</sup> | 29.1 (5) | 30.2 (5) | 27.8 (4) | 0.11 |
| VIS 1, n (%) | 19 (54.3) | 10 (52.6) | 9 (56.3) | >0.99 |
| Surgical interventions, n |  |  |  |  |
| Total knee arthroplasty | 23 | 11 | 12 |  |
| Exchange of total knee arthroplasty | 6 | 5 | 1 |  |
| Unicondylar sled prostheses | 4 | 2 | 2 |  |
| Open Reduction and Internal Fixation | 2 | 1 | 1 |  |

Abbreviations: BMI, body mass index; VIS, Visibility Score
a: p-value compares facet needle group versus tuohy needle group
b: chi-square used for comparison of categorical data, t-test for independent samples with variance homogeneity and Mann Whitney U-test for non parametric variables

**Figure 2.**
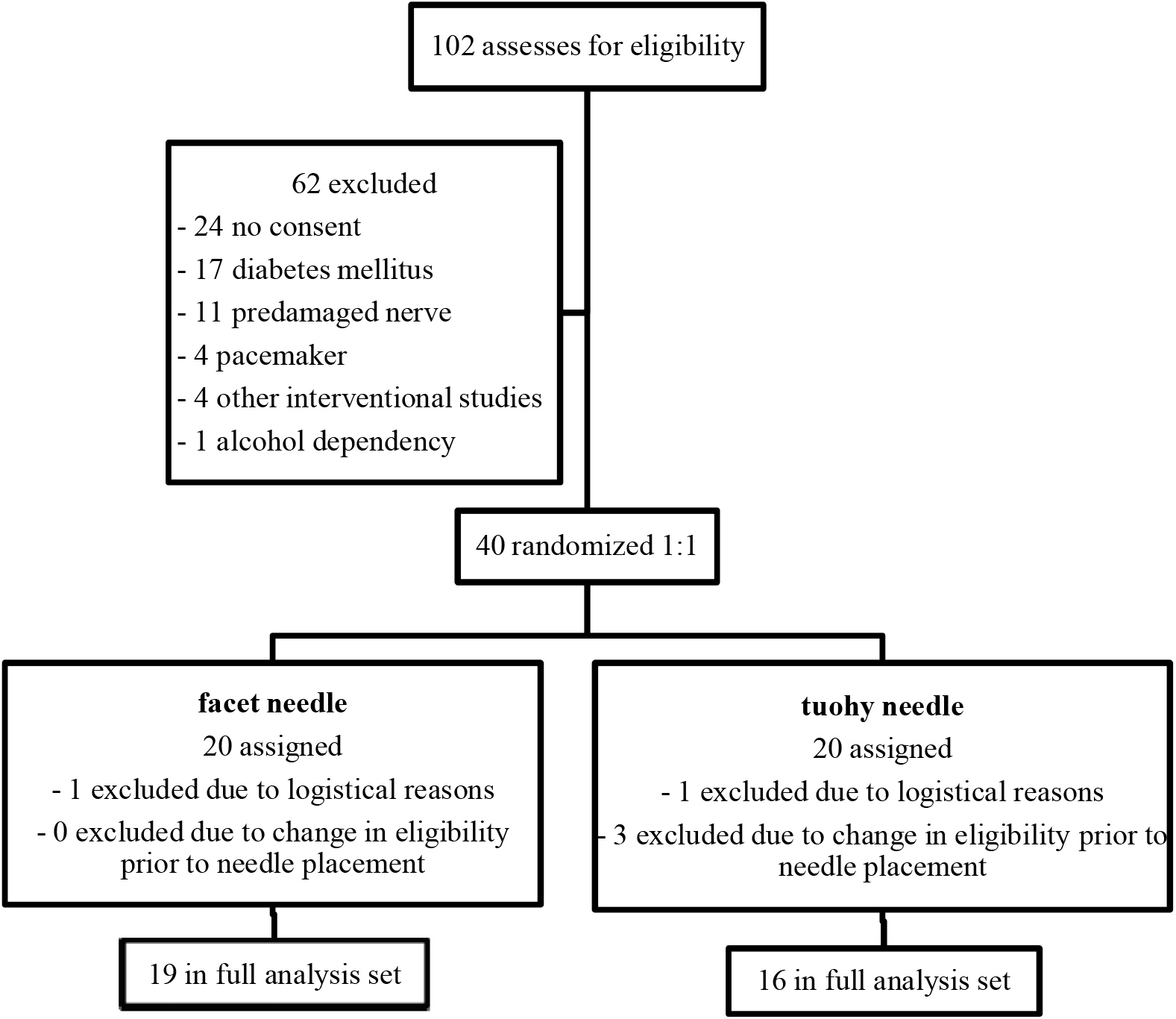
Study flowchart of full analysis set

### Stimulation success

The sciatic nerve was successfully stimulated in 23 of 35 trials. In 20 of these 23, an ultrasound verified contact between needle and nerve was necessary to provoke a motor response. In 3 cases a minimal distance between needle tip and nerve was assumed but could not be measured technically. In 56% of successful stimulations the initial current of 2 mA could not be reduced without losing stimulation response. In 8 cases the current could be reduced to 1 mA and in 2 participants a current of 0.5 mA resulted in a motor response at the sciatic nerve. The proportion of successful stimulations at direct contact between needle and nerve was significantly greater in facet group as displayed in figure 3. In 84.2% a motor response could be stimulated in facet group (N=16/19) whereas in tuohy group only 7 of 16 stimulations were successful (43.8%; p=0.03). Paresthesia was recorded in 6 participants in facet group and 5 in tuohy group. To improve the discrimination of structures in sonographic images hydrodissection with 5% glucose was used. In 3 cases ultrasound revealed a thin hyperechogenic tissue layer which separates from sciatic nerve during the injection of glucose. If this layer was penetrated, a prior missing motor response was generated. Only fractions of millimeters of needle movement decided about success or absence of stimulation response.

**Figure 3.**
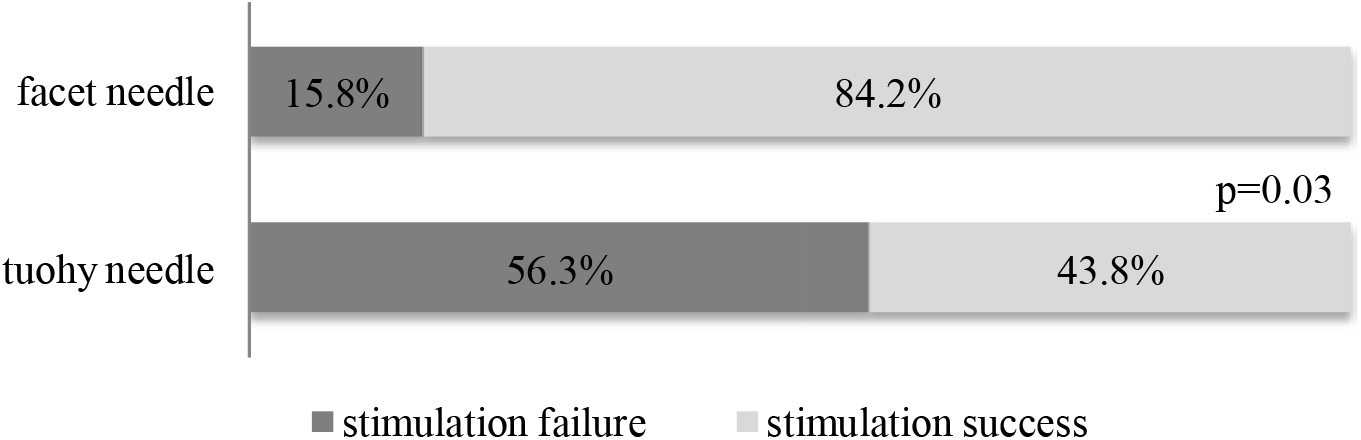
Proportion of stimulating success in study groups

### Secondary outcomes

The overall success rate of sensory block in recovery room was 62.9% (22/35) without statistically significant difference between groups (p=0.50). Among both groups participants with successful PNS had a significantly higher rate in efficient sensory nerve blockades in recovery room (Figure 4). If motor response could be triggered the success rate of sensory nerve block was 78.3% (18/23) compared with 33.3% (4/12; p=0.024) in patients without motor response to stimulation. No serious complication of regional anesthesia was recorded with one minor bleeding from puncture site.

**Figure 4.**
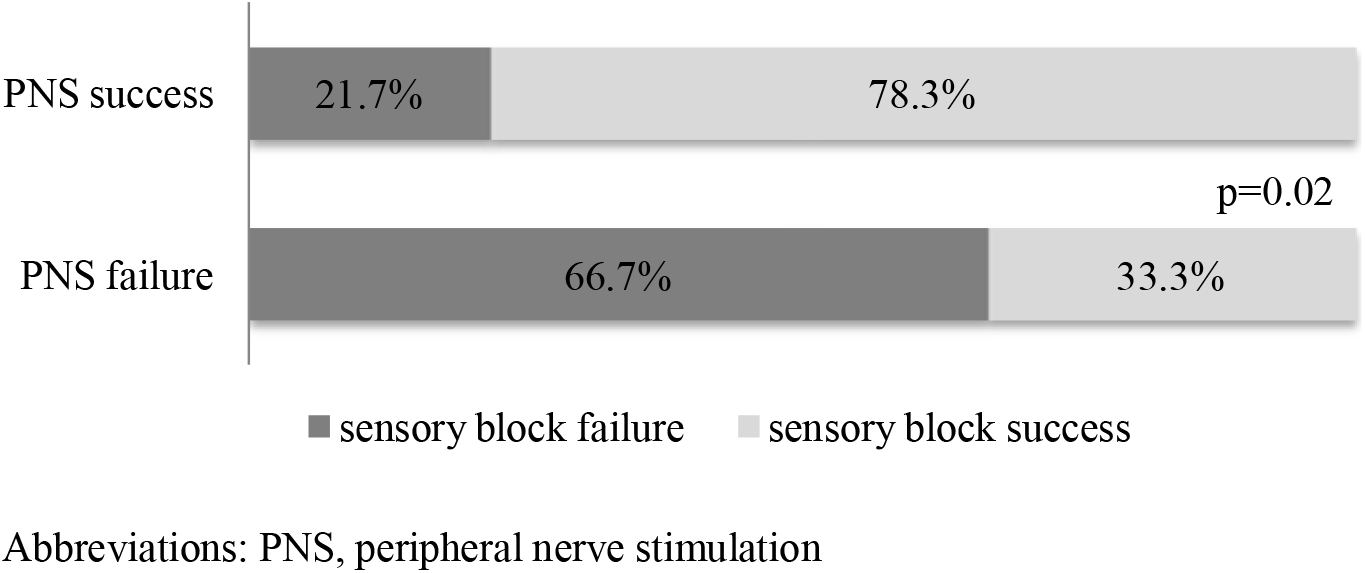
Proportion of successful nerve blockades in recovery room depending on stimulating success

### Age-adjusted analysis

We further explored influence of age on primary and secondary outcomes as study groups differed significantly (sensitivity analyses). However, the stimulation success (p=0.959), sensory block success (p=0.987) and visibility score (p=0.707) appeared unrelated with age per se. In a multivariate logistic regression model, age did not affect the stimulation success during block (adjusted OR 1.031, CI 0.956-1.111, p=0.426). Facet needle type was associated with an age-adjusted 9-fold increased probability of stimulation success (OR 9.158, 95% CI 1.524-55.026, p=0.02), thus validating abovementioned univariate findings (Hosmer-Lemeshow-test p=0.190 indicated sufficient model quality).

## DISCUSSION

This clinical pilot trial suggests that stimulation needles with small electrodes like the Uniplex needle with facet cut may provide greater stimulation success in proximal sciatic nerve block than tuohy needles with large electrodes. Due to the superior sensory block results after successful stimulation these data appear to be particularly relevant for the safety and success of the procedure.

The stimulation properties of needles observed in this study are comparable with those in a preclinical analysis. Chantrell et al. presented a model, which supports that the tuohy geometry and their insulation pattern greatly affect the propagation of electrical fields around the needle.^16^ The electric field intensity decreases with a larger surface area of the electrode and a constant electrical current. Therefore, needles with a smaller electrode surface at the needle tip are theoretically more likely to elicit an action potential when placed in the same location in relation to the nerve.

Nevertheless we observed several stimulation failures not just with touhy needles but also with facet needles. Other studies suggest that even in cases of intraneural placement there can be a stimulation failure.^10; 17^ This indicates that missing motor response does not exclude an adequate needle placement. In addition to that there is data supporting the hypothesis that the intensity of the current needed for PNS can vary due to the properties of the specific nerve and its surrounding tissue.^18; 19^ In this study targeting the sciatic nerve, more than half of the successful stimulations required a high current of 2 mA.

Andersen et al. described the tissue layers around the sciatic nerve including a paraneural sheath with a subparaneural space and suspected that these might influence regional anesthesia.^12^ Several studies discuss the paraneural sheath as a determining barrier for block success rates and time to block onset.^20^ The subparaneural space surrounding the epineurium of the sciatic nerve seems to be the most effective position for the injection of local anesthetics.^12; 13; 21^ Separation of a hyperechogenic membrane, suspected to be the paraneural sheath, during hydrodissection greatly improved the stimulation success in our investigation. Therefore it seems as if this layer not only affects the distribution of local anesthetics, but also success of PNS. It could be necessary to go into the subparaneural space carefully using hydrodissection and dual guidance technique to be able to override the isolating effects of the nonneural tissue. Thus an adequate stimulation response could be triggered and local anesthetics would be deposited closer to the nerve. In our study discrimination between the epineural and paraneural layer was difficult due to the resolution of the ultrasound. In order to prevent intraneural needle placement there might be cases with stimulation and injection above the paraneural sheath. This may be a reason of the 37% of sensory block failure at the recovery room in our study. Subparaneural injection of local anesthetics must therefore be distinguished from intraneural injection (subepineural or intrafascicular) with the potential risk of nerve lesion.^22^ Thus, the aim should be to inject the local anesthetic subparaneurally.

Another reason for varying block success among different studies might be the distribution of local anesthetics. Brull et al. presented a similar success rate of sensory block in US-guided popliteal sciatic block using a single-location technique which could be significantly increased to 94% by circumferential injection technique.^23^

Although facet needles with small electrodes show a significantly better stimulation success, which was associated with a successful sensory blockade, there was no significant correlation between needle types and block success. The missing correlation between successful PNS and success of motor block in contrast to sensory block may be explained by the small number of participants.

This study has potential limitations. Although there were significant differences in stimulation success between the compared groups the overall number of participants remains relatively small. As a pilot study it presents a basis for further studies and sample calculations. All conclusions based on these data allow only statements about PNS of the proximal sciatic nerve and just compare the two tested needles. Further an increased resolution of ultrasound imaging could improve the exact discrimination and measurement of needle tip placement.

Even though the survey followed a standardized procedure, the peripheral nerve blockade remains to be a procedure dependent upon the individual physician. Therefore a double blinded study design seems nearly unrealizable. A standardized method for outcome assessment by the same researcher was used to counteract these biases. Due to differences in age between the two groups, a subgroup analysis was performed to rule out the age of participants as an influencing factor on stimulation response.

## Conclusion

For a more reliable detection of contact between needle tip and nerve, using PNS for proximal sciatic nerve blocks, facet needles with a small electrode may be superior.

Further research is necessary to determine the influence of subparaneural needle placement on stimulation success. Additionally it could be investigated whether these findings can be transferred to PNS in peripheral nerves without a paraneural layer.

## Data Availability

All data produced in the present study are available upon reasonable request to the authors.

